# The indirect effect of the psychosocial work environment on the association between precarious employment and the production of steroid hormones: A cross sectional analysis

**DOI:** 10.1101/2022.01.30.22270116

**Authors:** Fabrizio Méndez Rivero, Oscar J Pozo, Mireia Julià

**Author notes:** **Funding** The research that gives rise to the results presented in this publication received funds from the National Research and Innovation Agency (ANII) of Uruguay under the code POS_EXT_2018_1_153741, and from the Spanish Ministry of Science and Innovation under grant agreement Nº CSO2017-89719-R (AEI/FEDER, UE).

## Abstract

**Objectives:** The main objectives of this article are (i) to explore the potential relationship between precarious employment and the production of steroid hormones (both adrenal and gonadal) and (ii) to evaluate the psychosocial risk factors at work (i.e. demands, control, and support) and work-life conflicts in this relationship.

**Methods:** Cross-sectional data were derived from a sample of workers from Barcelona (n=255 —125 men, 130 women). A set of 23 markers were determined from hair samples to evaluate the chronic production of both adrenal and gonadal steroids. Linear regression models were used to estimate the association between precarious employment and the production of adrenal and gonadal steroids, and decomposition analyses were applied to estimate the indirect effect of psychosocial risk factors and work-life conflict on this relationship.

**Results:** Gender differences in the association with PE-steroid production were found. Among men, gonadal axis steroids were associated with precarious employment (specifically, androstenedione and testosterone), while among women, adrenal axis steroids, primarily cortisol and markers derived from its metabolism, were associated with precarious employment. Psychosocial risk factors and work-life conflicts had significant positive indirect effects only among women.

**Conclusions:** Gender differences were found in respect of the indirect effects of psychosocial risk factors and work-life conflicts on the association between precarious employment and the production of adrenal and gonadal steroids, which suggests that, beyond the biochemical differences, the physiological effect of PE could be mediated by the social construction of gender identities, positions and roles in society and family.

**KEY MESSAGES:** 

**What is already known about this subject?:** Previous studies suggest that precarious employment is associated with workers’ health; however, most studies are based on self-rated health indicators and do not explore the causal mechanisms behind this association.

**What are the new findings?:** Precarious employment was associated with the production of some adrenal and gonadal steroids, and the psychosocial work environment had an indirect effect on this association, although with significant gender differences.

**How might this impact on policy or clinical practice in the foreseeable future?:** An occupational health policy aimed at improving the quality of employment and, at the same time, the psychosocial work environment can reduce the production of hormones that are associated with stress.

## BACKGROUND

Precarious employment (PE) refers to a generalized phenomenon of employment insecurity, income inadequacy, and a lack of rights and protection, which has been widely extended in recent decades in Europe and is recognized as a significant social determinant of health. Recently, after decades of studies based on one-dimensional proxies (e.g. temporary employment or perceived job insecurity), some multidimensional PE measurement instruments have been developed. One of them, the Employment Precariousness Scale (EPRES), has been highlighted as an insightful tool for operationalizing PE [1,2] through an instrument that encompasses six dimensions: temporariness, disempowerment, vulnerability, wages, rights, and the exercise of rights. [3].

Although several studies have shown that PE negatively affects working people’s physical and mental health, mechanisms and pathways linking PE to poor health outcomes have not been sufficiently explored. Some conceptual frameworks have been drawn up suggesting that PE could affect health through the psychosocial work environment. [4] From this approach, we argue that the psychosocial work environment is an intermediate step in a causal pathway that links economic, social, and political structures with health and disease through psychological and psychophysiological processes. [5] Work-life conflicts (WLC) have been also highlighted as a relevant psychological stressor in contemporary working life, which has increased amongst employees in most economic sectors, [6,7] especially women who have greater difficulties in reconciling the domestic sphere with paid work. [8]

The biological response to the psychosocial work environment means that it could be embodied - “get under the skin” - by workers altering their cognition, emotions, behavior, and physiology [5]. From the embodiment perspective, psychosocial working conditions could be embodied while bypassing the individual’s consciousness, [9] which implies that the embodiment of PRF should be measured through markers and not self-reported indicators. Hair cortisol has been used as indicator of damaging psychosocial work environments. The association between shiftwork, increased hair cortisol levels and abdominal obesity (a typical tissue effect of cortisol) has been found. [10] Unemployment is a constant stressor since it is connected to financial strain and psychological problems, and has also been associated with increased concentrations of hair cortisol. [11] Studies with ERI models show an association with job insecurity and higher levels of hair cortisol concentrations, [12] as well as suggesting prospective associations between ERI and cortisol. [13] However, to the best of our knowledge there are no studies on the indirect effect of PRFs on the relationship between multidimensional PE and hair cortisol. On the other hand, the use of hair cortisol as a biomarker of chronic stress has provided contradictory results. Whereas some studies proposed a positive correlation between hair cortisol levels and subjective measures of chronic stress, [14] others found either a poor correlation or none at all. [15-17] Cases of poor correlation may be partially due to the potential effect of cofounding factors such as age and body mass index. [18,19] The use of additional markers such as cortisol metabolites to evaluate the hypothalamic-pituitary-adrenal (HPA) and hypothalamic-pituitary-gonadal (HPG) axes can provide new insights into the relationship between HPA/HPG axis-employment and working conditions.

This study analyzes the relationship between PE and the production of steroid hormones (both adrenal and gonadal) in salaried workers from Barcelona city, as well as the indirect effects of a set of PRFs at work (i.e. psychological demands, control, and social support) and WLC on this relationship.

## MATERIALS AND METHODS

A cross-sectional study was conducted, based on a sample of 255 workers from Barcelona, Spain, aged 25-60 (125 men and 130 women). Further details of the sampling design may be found elsewhere. [20]

### Outcome variables

A comprehensive steroid profile was measured in hair based on a previously validated method. [21] Steroids and metabolites were divided between adrenal steroids (providing information about the HPA axis), including hair cortisol level 20α-dihydrocortisol (20αDHF), 20ß-dihydrocortisol (20βDHF), Cortisone, 20α-dihydrocortisone (20αDHE), 20ß dihydrocortisone (20βDHE), Cortolone, 11-dehydrocorticosterone and androstenedione (AED) and gonadal steroids (providing information about the HPG axis), also including AED, testosterone, and progesterone levels. Besides the hair concentrations of the targeted steroids, several ratios were included in order to evaluate the activity of key enzymes in the production and metabolism of steroids. As an example, the cortisol/cortisone ratio was calculated to evaluate the activity of the enzyme 11ß-hydroxysteroid dehydrogenase (responsible for the interconversion between cortisol and cortisone). Additionally, the relative abundance of each glucocorticosteroid (in %) was calculated as an additional marker. Since the distributions of the steroids and ratios are very dissimilar, the natural logarithm has been used to fit them to a normal distribution and obtain more reliable statistics.

### Explanatory variables

PE was measured through an adaptation of the EPRES validated to the PRESSED data (Supplementary file, ST1, ST2, ST3 shows psychometric properties). The scale consists of 24 indicators sorted into the EPRES’s dimensions specified above and another dimension related to extra working hours. Each dimension contributed equally to the total score, regardless of its number of items. To obtain an equal weight scale, each dimension score was computed independently, standardized, and integrated into a global summary score. Accordingly, the items in each dimension were added together, and the overall score was transformed into a 0 to 4 score. Then these scores were averaged into a global EPRES score, which ranged from 0 (not precarious) to 4 (most precarious). [3]

### Mediators

The WLC and PRFs dimensions “psychological demands”, “control”, and “social support” were measured using 32 items from the COPSOQ III. [22] Scores for each dimension were computed through simple averages of its corresponding items. Exploratory, confirmatory factor analyses and Cronbach’s alpha coefficients were used to evaluate the scales’ validity and reliability respectively (Supplementary material, ST4, ST5, ST6). Regarding the validity, factor-loading estimates revealed that all items were related to their theorized dimensions.

### Control variables

The covariates used for adjustment were age, body mass index (BMI), occupational social class (i.e., “Manual”, “Non-manual”), and a proxy of care work (people younger than 14 years old at home).

### Statistical analysis

A description of the studied sample was performed. Means and their standard deviation were calculated for continuous variables and prevalence and 95% CI for categorical variables (Table 1).

**Table 1.** Characteristics of the study population stratified by sex. Precarious Employment and Stress Study sample, 2020. [CI95%=95% confidence interval]

|  | Men<br>N=125 |  |  | Women<br>N=130 |  |  | p-<br>value |
| --- | --- | --- | --- | --- | --- | --- | --- |
|  | Mean | 95%CI |  | Mean | 95%CI |  |  |
| Age | 41.68 | 39.95 | 43.41 | 42.75 | 41.06 | 44.45 | 0.38 |
| Body mass index (BMI) | 25.34 | 24.71 | 25.97 | 24.75 | 24.00 | 25.49 | 0.23 |
| Occupational social class |  |  |  |  |  |  |  |
| No manual | 71.20 | 63.19 | 79.21 | 76.15 | 68.76 | 83.54 |  |
| Manual | 28.80 | 20.79 | 36.81 | 23.85 | 16.46 | 31.24 | 0.37 |
| Precarious employment (EPRES) | 1.04 | 0.95 | 1.13 | 1.01 | 0.92 | 1.10 | 0.67 |
| Psychosocial risk factors |  |  |  |  |  |  |  |
| Demands | 0.41 | 0.38 | 0.45 | 0.47 | 0.44 | 0.50 | 0.02 |
| Control | 0.30 | 0.27 | 0.33 | 0.82 | 0.25 | 0.31 | 0.44 |
| Support | 0.29 | 0.25 | 0.32 | 0.28 | 0.24 | 0.32 | 0.80 |
| Work-life conflict (WLC) | 0.44 | 0.40 | 0.48 | 0.47 | 0.43 | 0.52 | 0.30 |
| Care work | 0.54 | 0.40 | 0.67 | 0.61 | 0.46 | 0.75 | 0.48 |
| Adrenal and gonadal steroids |  |  |  |  |  |  |  |
| Cortisol | 10.47 | 8.56 | 12.39 | 9.35 | 8.03 | 10.68 | 0.35 |
| 20αDHF | 0.93 | 0.74 | 1.11 | 0.98 | 0.81 | 1.16 | 0.65 |
| 20βDHF | 5.47 | 4.70 | 6.23 | 4.51 | 4.01 | 5.01 | 0.04 |
| 20αDHE | 9.81 | 8.48 | 11.15 | 9.05 | 7.88 | 10.22 | 0.40 |
| 20βDHE | 7.29 | 6.24 | 8.33 | 5.39 | 4.79 | 5.99 | 0.00 |
| Cortisone | 34.15 | 30.82 | 37.49 | 27.34 | 24.23 | 30.45 | 0.00 |
| Androstenedione (AED) | 5.59 | 4.98 | 6.19 | 4.09 | 3.56 | 4.62 | 0.00 |
| Dihydrocorticosterone | 2.96 | 2.66 | 3.27 | 2.56 | 2.28 | 2.85 | 0.06 |
| Cortolone | 8.83 | 8.12 | 9.54 | 7.25 | 6.72 | 7.78 | 0.00 |
| Testosterone | 2.07 | 1.65 | 2.49 | 3.34 | -1.25 | 7.94 | 0.59 |
| Progesterone | 267.57 | -40.38 | 575.52 | 29.11 | 23.04 | 35.18 | 0.13 |
| Cortisol_Cortisona | 0.15 | 0.14 | 0.17 | 0.20 | 0.18 | 0.21 | 0.00 |
| %Cortisol | 1.39 | 1.32 | 1.45 | 1.64 | 1.57 | 1.71 | 0.00 |
| %Cortisone | 13.23 | 11.91 | 14.56 | 13.02 | 11.42 | 14.61 | 0.84 |
| %20αDHF | 0.30 | 0.26 | 0.33 | 0.35 | 0.31 | 0.39 | 0.04 |
| %20βDHF | 14.14 | 13.01 | 15.27 | 15.91 | 14.85 | 16.98 | 0.03 |
| %20αDHE | 51.85 | 50.14 | 53.56 | 49.01 | 47.44 | 50.58 | 0.02 |
| %20βDHE | 1.20 | 1.08 | 1.33 | 1.57 | 1.42 | 1.71 | 0.00 |
| 20βDHF/Cortisol | 8.01 | 7.56 | 8.47 | 8.20 | 7.82 | 8.58 | 0.52 |
| 20αDHF/Cortisol | 14.25 | 13.58 | 14.91 | 15.67 | 14.89 | 16.45 | 0.01 |
| 20αDHF/20βDHF | 10.55 | 10.07 | 11.02 | 9.64 | 9.27 | 10.01 | 0.00 |
| 20αDHE/20βDHF | 0.09 | 0.08 | 0.10 | 0.11 | 0.10 | 0.12 | 0.06 |
| Cortisone/dihydrocorticosterone | 0.64 | 0.59 | 0.69 | 0.58 | 0.53 | 0.62 | 0.08 |

Linear regression models were fitted to estimate the association of PE, PRFs and WLC with steroid production. Two models were estimated. Model 1 (crude) was adjusted for age and BMI. Model 2 (adjusted) was further adjusted for occupational social class, care work, demands, control, social support, and WLC. Only the results for steroids that were significantly associated with exposure or some of the mediators are presented (Table 2). The Karlson-Holm-Breen (KHB) method was used to estimate the indirect effect of PRFs and WLC on the relationship between PE and markers. Two models were fitted. Model 1 included demands, control, and social support as mediator variables, while in model 2, WLC was added as a mediator. Both models were adjusted for age, BMI, occupational social class, and care work (Table 3). All the analyzes were stratified by sex, thus, in order to compare coefficients between both sexes, measurement invariance across sexes was assessed for PE scale and PRF scales (Table 2 and Table 5 Supplemental material).

**Table 2.**
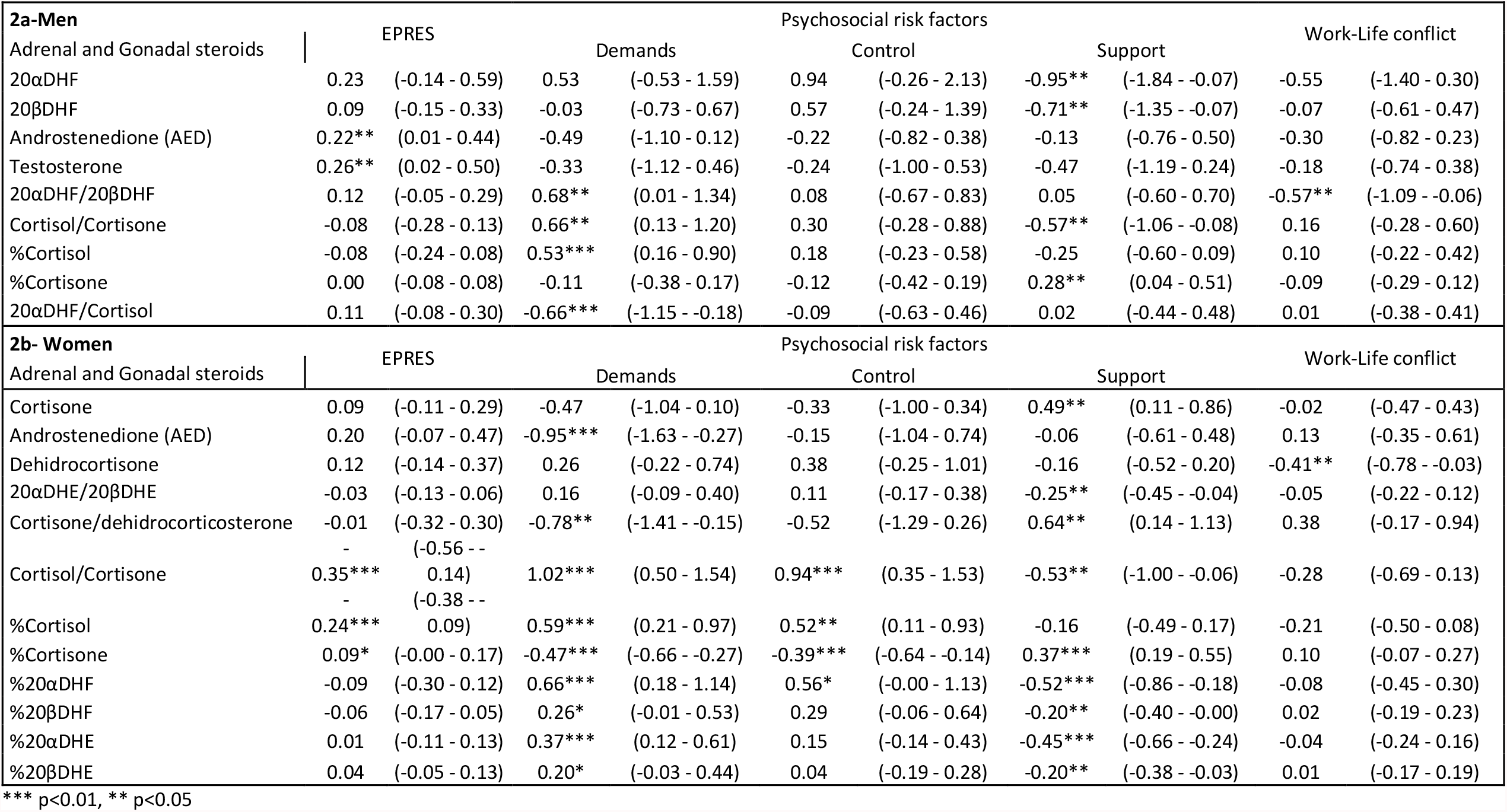
Linear regression coefficients and 95% confidence intervals (CI) for Production of Adrenal and Gonadal steroids and PE and PRFs concomitantly, adjusted for control variables and stratified by sex. Precarious Employment and Stress Study sample, 2020.

**Table 3.** Linear regression coefficients and 95% confidence intervals (CI) for the Production of Adrenal and Gonadal steroids and PE and PRFs, adjusted for control variables and stratified by sex, from the KHB-Method. Robust standard errors. Precarious Employment and Stress Study sample, 2020.

| **3a-Men** |  | **Direct** |  | **Total** |  | **Indirect** |  |
| --- | --- | --- | --- | --- | --- | --- | --- |
| **Model 1** |  | **Coeff.** | **95%CI** | **Coeff.** | **95%CI** | **Coeff.** | **95%CI** |
| Androstenedione (AED) |  | 0.13 | (-0.08 - 0.35) | 0.17 | (-0.04 - 0.39) | -0.04 | (-0.10 - 0.02) |
| Testosterone |  | 0.18 | (-0.05 - 0.40) | 0.23* | (-0.01 - 0.46) | -0.05 | (-0.13 - 0.02) |
| **Model 2** |  | **Coeff.** | **95%CI** | **Coeff.** | **95%CI** | **Coeff.** | **95%CI** |
| Androstenedione (AED) |  | 0.13 | (-0.07 - 0.34) | 0.22** | (0.00 - 0.44) | -0.09 | (-0.20 - 0.02) |
| Testosterone |  | 0.18 | (-0.05 - 0.40) | 0.26** | (0.02 - 0.49) | -0.08 | (-0.20 - 0.04) |
| **3a-Women** |  | **Direct** |  | **Total** |  | **Indirect** |  |
| **Model 1** |  | **Coeff.** | **95%CI** | **Coeff.** | **95%CI** | **Coeff.** | **95%CI** |
| Cortisol/Cortisone |  | -0.23** | (-0.40 - -0.05) | -0.39*** | (-0.59 - -0.20) | 0.17*** | (0.06 - 0.27) |
| %Cortisol |  | -0.17*** | (-0.30 - -0.04) | -0.27*** | (-0.41 - -0.14) | 0.10*** | (0.03 - 0.17) |
| **Model 2** |  | **Coeff.** | **95%CI** | **Coeff.** | **95%CI** | **Coeff.** | **95%CI** |
| Cortisol/Cortisone |  | -0.23*** | (-0.40 - -0.06) | -0.35*** | (-0.56 - -0.15) | 0.12** | (0.00 - 0.24) |
| %Cortisol |  | -0.17*** | (-0.29 - -0.05) | -0.24*** | (-0.38 - -0.10) | 0.07* | (-0.01 - 0.15) |

\*\*\* p<0.01, \*\* p<0.05

The KHB method allows the unbiased comparison of regression coefficients between models and the decomposition of mediation effects. [23,24]

## RESULTS

The characteristics of the sample studied are shown in Table 1. Gender differences on markers levels were significant for 20αDHF, 20αDHE, Cortisone, AED, Cortolone, Testosterone, %20αDHE, 20αDHF/Cortisol, which were higher among men; and Cortisol/Cortisone, %Cortisol, %20αDHF, %20βDHF, %20βDHE 20αDHF/20βDHF, which were higher among women. No significant gender differences were found for PE. Regarding PRFs, “Demands” was significantly higher among women (0.47; 95%CI: 0.44-0.50 vs. 0.41; 95%CI: 0.38-0.45). Gender differences for WLC were not significant.

The association of markers with PE and PRFs is presented in Table 2. Linear regression coefficients adjusted for control variables are shown in Table 2a for men and 2b for women. Among men, AED (β=0.22; 95%CI: 0.01-0.44) and Testosterone (β=0.26; 95%CI: 0.02-0.50) were associated with PE. Concerning PRFs, 20αDHF/20βDHF (β=0.68; 95%CI: 0.01 - 1.34), Cortisol/Cortisone (β=0.66; 95%CI: 0.13-1.20) and %Cortisol (β=0.53; 95%CI: 0.16 - 0.90) were positively associated with “Demands”, whilst 20αDHF/Cortisol (β=-0.66; 95%CI: -1.15 - -0.18) was negatively associated. There are no steroids associated with “Control”. “Social Support” showed positive coefficients for %Cortisone (β=0.28; 95%CI: 0.04 - 0.51), and negative for 20αDHF (β=-0.95; 95%CI: -1.84 - -0.07), 20βDHF(β=- 0.71; 95%CI: -1.35 - -0.07) and Cortisol/Cortisone (β=--0.57; 95%CI: -1.06 - -0.08). WLC was negatively associated with 20αDHF/20βDHF (β=-0.57; 95%CI:-1.09 - -0.06). Among women, negative associations between PE and Cortisol/Cortisone (β=-0.35; 95%CI: -0.56 - -0.14) and %Cortisol (β=-0.24; 95%CI: -0.38 - -0.09) were found. Concerning PRFs, “Demands” showed positive coefficients for Cortisol/Cortisone (β=1.02; 95%CI: 0.50 - 1.54), %Cortisol (β=0.59; 95%CI: 0.21 - 0.97), %20αDHF (β=0.66; 95%CI: 0.18 - 1.14), %20αDHE (β=0.37; 95%CI: 0.12 - 0.61), and negative coefficients for AED (β=-0.95; 95%CI: -1.63 - -0.27), Cortisone/dehidrocorticosterone (β=-0.78; 95%CI: -1.41 - -0.15), and %Cortisone (β=-0.47; 95%CI: -0.66 - -0.27). “Control” was positively associated with Cortisol/Cortisone (β=0.94; 95%CI: 0.35 - 1.53) and %Cortisol (β=0.52; 95%CI: 0.11 - 0.93), and negatively associated with %Cortisone (β=-0.39; 95%CI: -0.64 - -0.14). Cortisone (β=0.49; 95%CI:0.11 - 0.86), Cortisone/dehidrocorticosterone (β=0.64; 95%CI: 0.14-1.13) and %Cortisone (β=0.37; 95%CI:0.19-0.55) were positively associated with “Social Support”, whilst 20αDHE/20βDHE (β=-0.25; 95%CI: -0.45- -0.04), Cortisol/Cortisone (β=-0.53; 95%CI: -1.00 - -0.06), %20αDHF (β=-0.52; 95%CI: -0.86 - - 0.18), %20βDHF (β=-0.20; 95%CI: -0.40 - -0.00), %20αDHE (β=-0.45; 95%CI: -0.66 - -0.24) and %20βDHE (β=-0.20; 95%CI: -0.38 - -0.03) were negatively associated. WLC showed a negative coefficient for Dehidrocortisone (β=-0.41; 95%CI:-0.78 - -0.03).

Table 3 shows the results of KHB decomposition analyses for those steroids that were associated with PE. Thus, the indirect effect of PE through the PRFs and work-life conflict could be estimated, while the comparison between models allows estimating the change in the indirect effect when work-life conflict is added as a mediating variable. Among men, no indirect effect of PRFs was observed. There were no significant indirect effects when adding WLC as a mediator, although a significant total effect was observed in both steroids ((β_AED_: 0.22; 95% CI: 0.02 - 0.43)) ; (β_Testosterone_: 0.26; 95% CI: 0.02 - 0.50)). Among women, significant indirect effects for Cortisol (β= 0.18; 95% CI: 0.04 - 0.32), Cortisol/Cortisone (β 0.19; 95% CI: 0.08 - 0.31) and %Cortisol (β 0.12; 95% CI: 0.05 - 0.20) were found in Model 1, while, when incorporating WLC as a mediator, the indirect effect for Cortisol was not significant, and the magnitude of the effect decreased for the other steroids ((β_Cortisol/Cortisone_: 0.15; 95% CI:0.02 - 0.28) ; (β_% Cortisol_: 0.09; 95% CI:0.01 - 0.18)).

## DISCUSSION

The main objectives of this article were (i) to explore the potential relationship between PE and the production of steroid hormones (both adrenal and gonadal); and (ii) to evaluate the psychosocial work environment as a possible mediator in such a relationship in a sample of salaried workers from Barcelona city, both men and women. The main results suggest the existence of a relationship between PE and the production of steroid hormones, adjusted for PRFs and WLC. Remarkably, it has found a gender difference in that relationship. A significant positive association between androgens, i.e., gonadal steroids, (AED and testosterone) and PE was found among men. In contrast, women showed a negative association between EP and corticosteroids, i.e., adrenal steroids (cortisol and metabolites). Several potential explanations might lie behind these results.

From a biochemical point of view, gender differences in the production of steroid hormones and their relationship with stress have been previously described. [25] Our results suggest that PE would increase the production of gonadal steroids among men, leading to a subsequent rise in aggressiveness and dominant behaviour, [26] and thus pointing out the pivotal role of the HPG axis in men. In contrast, our results suggest that the role of the HPA axis is more important for women. The key function of the HPA axis in the relationship between PE and steroids is not surprising since overproduction of cortisol is a common biological feature under stress conditions. More surprising is the negative association observed between PE and several metabolites related to the HPA axis. Although several studies have shown gender differences in cortisol production after stressful events, [27] it is difficult to believe that the negative correlation is exclusively due to biochemical reasons.

Sociological factors such as working conditions might also exacerbate gender differences in response to stressful situations. Previous studies have found gender differences in occupational health related to working conditions linked to structural gender inequality in labour markets. [28] A preliminary hypothesis could be that the physiological effect of PE could be mediated by the social construction of gender identities, differentially affecting men and women. In a patriarchal system marked by the sexual division of work, in which the masculine role mainly draws on the “male breadwinner” stereotype, men can be psychologically affected by the perception of not meeting the social expectations associated with their role. Thus, the increased production of gonadal steroids could be a way for men to respond psycho-physiologically to PE. This hypothesis is based on classic social psychology approaches suggesting that the impact of employment problems on health is related to the different positions and roles available for men and women in society and the family.[29] For example, it has been found that unemployment was more negatively related to mental health among men than among women in a gender regime in which the need for employment differs between the sexes (Ireland), while men and women were equally affected by unemployment in a gender regime with a similar need for employment (Sweden). [30]

Therefore, men and women have different psychosocial and economic employment needs based on gender roles. [31] In fact, in this study found that, among women, the association of some steroids with PE presents negative coefficients, showing an inverse relationship to that hypothesised, maybe because, unlike men, women’s perceptions of PE are not mediated by the role of providers. Furthermore, the position of women in the sexual division of labour as the main partner responsible for care and home duties may imply that some characteristics of PE, such as flexibility or a low workload, are perceived as beneficial because they contribute to reconciling paid and unpaid work. [32]

It should be noted that, although the sexual division of work has been losing its rigidity over time, mainly due to the massive and sustained entry of women into the job market, there has not been any effective redistribution of responsibilities within the family, where changes are slower and co-responsibility between men and women is still a long way off. [33,34]. Besides, gender relations within a family framework still tend to be patriarchal, and even if occupational status is higher, women rarely have enough power to force men to agree to an equitable division of domestic work and childcare. [35] Regarding the psychosocial work environment, it was found that, for both men and women, high demands and low social support are the two psychosocial factors associated with the production of the highest number of steroids. Although the meaning of these associations is not entirely conclusive, it is noteworthy that for low social support, the associations with most steroids are negative, while for high demands the majority are positive. This implies that, while low social support increases steroid production, high demands reduce it, suggesting that the latter could be a protective mechanism. In this sense, several previous studies of mental health have found that high demands reduce the risk of depression and anxiety disorders. [36, 37]

One of the main objectives of this study was to estimate the indirect effect of PRFs and WLC on the relationship between PE and steroid production. The existence of significant indirect effects would indicate that a proportion of the association of the exposure and the outcome of interest crystallizes through the psychosocial work environment. The results show significant indirect effects only for women, suggesting the existence of gender differences in the psychophysiological response to PE. A recent study found a full mediation of PRF on PE and mental health among women and a partial mediation among men. The authors suggest that women are more exposed to worse working conditions, including the psychosocial environment, due to occupational segregation of gender in the labour market.[38] Both results show that the psychosocial work environment has a greater weight in women’s psychological and physiological responses to PE than with men. Thus, women may react more to proximal factors such as the psychosocial environment than to distal factors such as PE, while precisely the opposite occurs among men. The study does not allow further progress on the possible causes of these differences. However, it will be necessary to delve into gender differences in perceptions of working and employment conditions in the future.

### Strength and limitations

This study has some of the limitations that are inherent in a cross-sectional design. Firstly, it is not possible to extract a direct causal effect, and a possible reverse causality must be considered: a high production of steroids (which could indicate psycho- physiological alterations) at the beginning of the study may increase the chances of having a precarious job or an unfavorable psychosocial environment. Second, there is no information on the period during which these individuals have been exposed to PE or PRF, which may somewhat alterhe results. Therefore, further longitudinal studies are needed.

On the other hand, a notable strength of this article is its use of biological markers, something new not just in the study of PE, but also in the field of social epidemiology, where subjective and/or self-reported health measures are usually used. In turn, in biochemical research, simultaneously studying the two axes (gonadal and adrenal) in hair is also new. In previous studies, only steroids of the gonadal axis have been studied. Furthermore, this article shows the importance of taking employment conditions into account in the study of psychosocial working conditions. Most psychosocial risk models theoretically assume social causality, where the organization of work determines the psychosocial work environment, but they do not explain the individual’s relationship with the environment. [39] Furthermore, assimilating the “social” to the “psychological” means that the models are unable to explain how the social structure determines the psychosocial work environment. [40] Taking the PE into account allows us to explain how the political context and labour relations determine the organization of work in a complex process that impacts on workers’ health. Therefore, empirical advances such as those offered in this article stimulate the development of new theoretical and methodological frameworks that relate the psychosocial risks to PE to explain the global impact of the workplace on health.

#### Concluding remarks

Gender differences were found in the association between the production of PE steroids (both adrenal and gonadal) and the indirect effects of PRF and WLC. Biochemically, this could indicate the pivotal role of the HPG axis in men, the HPA axis being more important for women. In turn, these results suggest that the physiological effect of PE could be mediated by the social construction of gender identities that draw on the “male breadwinner” stereotype. This contributes to supporting the hypothesis that the influence of PE on health is related to the different positions and roles of men and women in society and the family. Future studies should delve further into these differences in the relationship between precarious employment, PRFs and their psycho-physiological effect to improve employment and working policies, especially from the perspective of the social determinants of health.

## Data Availability

All data produced in the present study are available upon reasonable request to the authors.

## ACKNOWLEDGMENTS

The authors would like to thank Mireia Bolíbar, who, as principal investigator of the project giving rise to these results, made a fundamental contribution to the conception and design of the article. The authors also thanks Alex Gómez-Gómez for his important contribution to this article by analyzing the hormones in the hair samples.

(In Title page to avoid the identification of some authors)

