## Supplemental Tables for "The indirect effect of the psychosocial work environment on the association between precarious employment and the production of steroid hormones: A cross sectional analysis"

Supplementary Table S1. Goodness-of-ft of EPRES scale from exploratory factor analysis. Precarious Employment and Stress Study sample, 2020 , N men=7650, N women= 8282.

| | Factor1 | Factor2 | Factor3 | Factor4 | Factor5 | Factor6 | Factor7 | Uniqueness |
|---|---|---|---|---|---|---|---|---|
| Type of contract | | | 0.26 | 0.62 | -0.16 | | 0.23 | 0.47 |
| How long in total have you been working for this company? | | | 0.30 | 0.34 | -0.15 | | | 0.75 |
| Approximately how much do you earn net per month? | 0.21 | | 0.66 | 0.11 | -0.16 | | | 0.49 |
| How often does your current salary allow you to cover your basic daily needs? | 0.10 | | 0.73 | | | | | 0.43 |
| How often does your current salary allow you to cover major unforeseen expenses? | | 0.16 | 0.76 | 0.11 | | | | 0.38 |
| How were the working conditions in relation to your working hours decided? | | 0.11 | | | | 0.69 | | 0.51 |
| How were the working conditions in relation to your salary decided? | | | | | | 0.67 | | 0.54 |
| How often in this company are you afraid to demand better working conditions? | 0.20 | 0.48 | | | | | | 0.72 |
| How often in this company do you feel defenceless against unfair treatment by your superiors? | 0.24 | 0.67 | | | | 0.12 | | 0.47 |
| How often in this company would you be fired from your job if you did not do everything they asked you to do? | | 0.34 | 0.15 | | | | 0.28 | 0.77 |
| How often in this company are you treated in an authoritarian way? | 0.21 | 0.70 | | | | | | 0.45 |
| How often in this company do they make you feel that you can easily be replaced? | 0.11 | 0.69 | 0.18 | | | | | 0.48 |
| Entitlement to paternity/paternity leave. | 0.10 | | 0.22 | 0.48 | | | -0.28 | 0.62 |
| Entitlement to retirement pension, disability. | 0.21 | | 0.40 | 0.34 | | | -0.26 | 0.61 |
| Entitlement to unemployment benefit | | | | 0.46 | 0.11 | 0.13 | 0.13 | 0.74 |
| Do you have the right to severance pay? | | | | 0.61 | | | -0.13 | 0.61 |
| In this company you can take your weekly holidays without any problems | 0.20 | | | | | 0.19 | 0.22 | 0.86 |
| In this company, you can take your holidays without any problems. | 0.31 | 0.15 | 0.15 | 0.14 | | | 0.35 | 0.71 |
| In this company you can take a day off for family reasons (care of children, dependants, sick people, etc.) without any problem | 0.61 | 0.36 | | 0.19 | | | -0.10 | 0.44 |
| In this company you can take a day off for personal reasons without any problem | 0.68 | 0.31 | | | | | -0.13 | 0.41 |
| In this company you can take sick leave without any problem | 0.63 | | 0.18 | 0.15 | | 0.11 | 0.19 | 0.49 |

| | | | | | | | | |
|---|---|---|---|---|---|---|---|---|
| In this company, you can go to the doctor when you need to. | 0.71 | 0.12 | 0.11 | | | | | 0.46 |
| Voluntarily work overtime | | 0.10 | | | 0.75 | | | 0.41 |
| Number of overtime hours you work | | | | | 0.76 | | | 0.40 |
| Changes in working hours, If yes, how far in advance are you informed of such changes? | 0.15 | | 0.14 | | | 0.23 | -0.18 | 0.86 |

Supplementary Table S2. Goodness-of-ft of EPRES scale from confirmatory factor analysis. Precarious Employment and Stress Study sample, 2020, N men=125, N women= 130.

| | χ2 [df]. p-value | CFI | ΔCFI | TLI | ΔTLI | RMSEA (90% CI) | ΔRMSEA |
|---|---|---|---|---|---|---|---|
| Regular CFA | | | | | | | |
| | 322.909 [231]; p<0.001 | 0.989 | - | 0.987 | - | 0.040 (0.030-0.050) | - |
| Multi-group CFA by women and men | | | | | | | |
| Configural invariance | 628.514 [462]; p<0.001 | 0.982 | 0 | 0.979 | 0 | 0.054 (0.043-0.064) | 0.014 |
| Metric invariance | 659.604 [479]; p<0.001 | 0.981 | -0.001 | 0.978 | -0.001 | 0.055 (0.044-0.065) | 0.001 |
| Thresholds invariance | 683.268 [541]; p<0.001 | 0.985 | 0.004 | 0.985 | 0.007 | 0.046 (0.034-0.056) | -0.009 |

Supplementary Table S3. Reliability of EPRES scale from Cronbach´s alpha. Precarious Employment and Stress Study sample. 2020. N men=125. N women= 130.

| Item | Obs | Sign | Item-test correlation | Item-rest correlation | Average interitem covariance | Alpha |
|---|---|---|---|---|---|---|
| Salary | | | | | | |
| Approximately how much do you earn net per month? | 254 | + | 0.76 | 0.59 | 1.16 | 0.77 |
| How often does your current salary allow you to cover your basic daily needs? | 254 | + | 0.86 | 0.67 | 0.72 | 0.65 |
| How often does your current salary allow you to cover major unforeseen expenses? | 254 | + | 0.90 | 0.70 | 0.54 | 0.66 |
| Test scale | | | | | 0.80 | 0.78 |
| Vulnerability | | | | | | |
| How often in this company are you afraid to demand better working conditions? | 255 | + | 0.67 | 0.44 | 0.74 | 0.71 |
| How often in this company do you feel defenceless against unfair treatment by your superiors? | 255 | + | 0.74 | 0.57 | 0.68 | 0.66 |
| How often in this company would you be fired from your job if you did not do everything they asked you to do? | 255 | + | 0.62 | 0.33 | 0.81 | 0.76 |
| How often in this company are you treated in an authoritarian way? | 255 | + | 0.75 | 0.61 | 0.70 | 0.65 |
| How often in this company do they make you feel that you can easily be replaced? | 255 | + | 0.76 | 0.59 | 0.64 | 0.65 |
| Test scale | | | | | 0.71 | 0.73 |
| Rights | | | | | | |
| Entitlement to paternity/paternity leave. | 255 | + | 0.71 | 0.43 | 0.07 | 0.51 |
| Entitlement to retirement pension, disability. | 255 | + | 0.67 | 0.37 | 0.08 | 0.56 |
| Entitlement to unemployment benefit | 255 | + | 0.57 | 0.29 | 0.10 | 0.61 |
| Do you have the right to severance pay? | 255 | + | 0.75 | 0.48 | 0.06 | 0.46 |
| Test scale | | | | | 0.08 | 0.61 |
| Vulnerability | | | | | | |
| In this company you can take your weekly holidays without any problems | 255 | + | 0.44 | 0.28 | 0.67 | 0.79 |
| In this company, you can take your holidays without any problems. | 255 | + | 0.59 | 0.40 | 0.58 | 0.76 |
| In this company you can take a day off for family reasons (care of children, dependants, sick people, etc.) without any problem | 255 | + | 0.76 | 0.60 | 0.47 | 0.71 |
| In this company you can take a day off for personal reasons without any problem | 255 | + | 0.79 | 0.64 | 0.45 | 0.70 |
| In this company you can take sick leave without any problem | 255 | + | 0.72 | 0.56 | 0.50 | 0.73 |
| In this company, you can go to the doctor when you need to. | 255 | + | 0.74 | 0.60 | 0.50 | 0.72 |
| Test scale | | | | | 0.53 | 0.77 |
| Salary | | | | | | |
| Voluntarily work overtime | 253 | + | 0.77 | 0.46 | 0.19 | 0.19 |
| Number of overtime hours you work | 253 | + | 0.81 | 0.49 | 0.08 | 0.09 |

| | | | | | | |
|---|---|---|---|---|---|---|
| Changes in working hours, If yes, how far in advance are you informed of such changes? | 253 | + | 0.56 | 0.08 | 1.08 | 0.78 |
| Test scale | | | | | 0.45 | 0.51 |

Supplementary Table S4. Goodness-of-ft PRF´s and WLC scales from exploratory factor analysis. Precarious Employment and Stress Study sample, 2020. , N men=7650, N women= 8282.

| Variable | Factor 1 | Factor 2 | Factor 3 | Factor 4 | Uniqueness |
|---|---|---|---|---|---|
| How often do you not have time to complete all your work tasks? | 0.28 | -0.17 | 0.55 | | 0.59 |
| Do you have enough time for your work tasks? | 0.37 | | 0.53 | | 0.56 |
| Do you have to work very fast? | 0.40 | 0.12 | 0.43 | 0.12 | 0.63 |
| Do you have to deal with other people's personal problems as part of your work? | | | 0.49 | -0.12 | 0.74 |
| Do you work at a high pace throughout the day? | 0.35 | | 0.54 | | 0.58 |
| Is your work emotionally demanding? | 0.31 | 0.14 | 0.65 | | 0.46 |
| Does your work require that you hide your feelings? | 0.20 | 0.26 | 0.47 | | 0.66 |
| Do you have a large degree of influence on the decisions concerning your work? | | 0.20 | -0.13 | 0.65 | 0.52 |
| Do you have any influence on HOW you do your work? | | 0.17 | | 0.67 | 0.50 |
| Do you have the possibility of learning new things through your work? | 0.12 | 0.51 | -0.32 | 0.32 | 0.52 |
| Can you use your skills or expertise in your work? | | 0.45 | -0.30 | 0.32 | 0.60 |
| Do you feel that the work you do is important? | | 0.42 | | 0.13 | 0.80 |
| How often do you get help and support from your immediate superior, if needed? | 0.19 | 0.65 | 0.18 | 0.17 | 0.48 |
| How often does your immediate superior talk with you about how well you carry out your work? | | 0.63 | | 0.23 | 0.55 |
| How often do you get help and support from your colleagues, if needed? | | 0.64 | | | 0.58 |
| Do you feel part of a community at your place of work? | | 0.63 | | | 0.59 |
| Do you feel that your work drains so much of your energy that it has a negative effect on your private life? | 0.76 | 0.12 | 0.21 | | 0.35 |
| Do you feel that your work takes so much of your time that it has a negative effect on your private life? | 0.85 | | 0.10 | | 0.26 |
| Are there times when you need to be at work and at home at the same time? | 0.42 | | 0.15 | -0.18 | 0.77 |
| The demands of my work interfere with my private and family life? | 0.68 | | 0.16 | | 0.50 |

Supplementary Table S5. Goodness-of-ft PRF´s and WLC scales from confirmatory factor analysis. Precarious Employment and Stress Study sample, 2020, N men=125, N women= 130.

| | χ2 [df], p-value | CFI | ΔCFI | TLI | ΔTLI | RMSEA (90% CI) | ΔRMSEA |
|---|---|---|---|---|---|---|---|
| Regular CFA | | | | | | | |
| | 732.672 [203]; p<0.001 | 0.939 | - | 0.930 | - | 0.104 (0.096-0.112) | - |
| Multi-group CFA by women and men | | | | | | | |
| Configural invariance | 1044.355 [406]; p<0.001 | 0.935 | 0 | 0.926 | 0 | 0.114 (0.106-0.123) | 0.010 |
| Metric invariance | 10344.527 [462]; p<0.001 | 0.930 | -0.005 | 0.924 | -0.002 | 0.116 (0.108-0.124) | 0.002 |
| Thresholds invariance | 31.217.153 [368]; p<0.001 | 0.935 | 0.005 | 0.938 | -0.014 | 0.105 (0.097-0.113) | -0.011 |

Supplementary Table S6. Reliability of PRF`s and WLC scales from Cronbach´s alpha. Precarious Employment and Stress Study sample. 2020. N men=125. N women= 130.

| Item | Obs | Sign | Item-test correlation | Item-rest correlation | Average interitem covariance | Alpha |
|---|---|---|---|---|---|---|
| Demands | | | | | | |
| How often do you not have time to complete all your work tasks? | 254 | + | 0.64 | 0.48 | 0.03 | 0.76 |
| Do you have enough time for your work tasks? | 254 | + | 0.65 | 0.51 | 0.03 | 0.75 |
| Do you have to work very fast? | 254 | + | 0.61 | 0.48 | 0.04 | 0.76 |
| Do you have to deal with other people's personal problems as part of your work? | 254 | + | 0.60 | 0.41 | 0.04 | 0.77 |
| Do you work at a high pace throughout the day? | 254 | + | 0.69 | 0.57 | 0.03 | 0.74 |
| Is your work emotionally demanding? | 254 | + | 0.77 | 0.64 | 0.03 | 0.72 |
| Does your work require that you hide your feelings? | 254 | + | 0.64 | 0.46 | 0.03 | 0.76 |
| Test scale | | | | | 0.03 | 0.78 |
| Control | | | | | | |
| Do you have a large degree of influence on the decisions concerning your work? | 255 | + | 0.70 | 0.48 | 0.03 | 0.66 |
| Do you have any influence on HOW you do your work? | 255 | + | 0.64 | 0.43 | 0.03 | 0.68 |
| Do you have the possibility of learning new things through your work? | 255 | + | 0.77 | 0.56 | 0.02 | 0.62 |
| Can you use your skills or expertise in your work? | 255 | + | 0.73 | 0.53 | 0.02 | 0.64 |
| Do you feel that the work you do is important? | 255 | + | 0.55 | 0.36 | 0.03 | 0.70 |
| Test scale | | | | | 0.03 | 0.71 |
| Support | | | | | | |
| How often do you get help and support from your immediate superior. if needed? | 246 | + | 0.81 | 0.63 | 0.03 | 0.66 |
| How often does your immediate superior talk with you about how well you carry out your work? | 246 | + | 0.78 | 0.56 | 0.03 | 0.71 |
| How often do you get help and support from your colleagues. if needed? | 246 | + | 0.73 | 0.54 | 0.04 | 0.72 |
| Do you feel part of a community at your place of work? | 246 | + | 0.72 | 0.52 | 0.04 | 0.72 |
| Test scale | | | | | 0.03 | 0.76 |
| Work-life conflict | | | | | | |
| Do you feel that your work drains so much of your energy that it has a negative effect on your private life? | 253 | + | 0.85 | 0.70 | 0.04 | 0.72 |
| Do you feel that your work takes so much of your time that it has a negative effect on your private life? | 253 | + | 0.89 | 0.76 | 0.04 | 0.68 |
| Are there times when you need to be at work and at home at the same time? | 253 | + | 0.61 | 0.40 | 0.07 | 0.85 |
| The demands of my work interfere with my private and family life? | 253 | + | 0.81 | 0.65 | 0.05 | 0.74 |
| Test scale | | | | | 0.05 | 0.81 |
